# Alterations to the gut microbiome after sport-related concussion and subconcussive impacts in a collegiate football players cohort

**DOI:** 10.1101/2021.07.10.21260292

**Authors:** Sirena Soriano, Kristen Curry, Saeed Sadrameli, Qi Wang, Michael Nute, Elizabeth Reeves, Rasadul Kabir, Jonathan Wiese, Amber Criswell, Sarah Schodrof, Gavin Britz, Rajan Gadhia, Kenneth Podell, Todd Treangen, Sonia Villapol

**Author notes:** **Correspondence and request for materials should be addressed to** Sonia Villapol, Ph.D. Current address; Department of Neurosurgery, Houston Methodist Research Institute, 6670 Bertner Avenue, R10-117, Houston, 77030, TX. These authors contributed equally to this work (Sirena Soriano, Kristen Curry, Saeed Sadrameli).

## Abstract

Concussions, both single and repetitive, during contact sports cause brain and body alterations in athletes. The role of the brain-gut connection and changes in the microbiota have not been well established after a head injury or concussion-related health consequences. We recruited 33 Division I Collegiate football players and collected blood, stool, and saliva samples throughout the athletic season. Analysis of the gut microbiome reveals a decrease in abundance for two bacterial species, *Eubacterium rectale*, and *Anaerostipes hadrus*, after a diagnosed concussion. No significant differences were found regarding the salivary microbiome. Serum biomarker analysis shows an increase in GFAP blood levels in athletes during athletic activity. Additionally, S100β and SAA blood levels were positively correlated with the abundance of *Eubacterium rectale* species among athletes exposed to subconcussive impacts. These novel findings provide evidence that detecting changes in the gut microbiome may pave the way for improved concussion diagnosis following head injury.

## INTRODUCTION

Concussions (or mild traumatic brain injury [mTBI]) affect 10-20% of athletes in collision sports^1^. Multiple occurrences of such events are referred to here as repetitive mTBI. A concussion is a complex pathophysiological process affecting the brain, induced by biomechanical forces ^2^. Sports-related concussion (SRC) is a form of mTBI common in collision sport athletes that has been linked to long-term neurological abnormalities ^3-5^. However, subconcussive impacts are defined as events similar to those giving rise to a concussion but involving insufficient impact forces or accelerations to produce symptoms associated with mTBI^6^.

In a typical season, athletes in collision sports often suffer both cumulative effects of repetitive subconcussive impacts and concussions. Due to the subjective nature of mTBI and the asymptomatic nature of sub-concussive head impacts, athletes frequently underreport symptoms and suffer repetitive blows after an injury resulting in implications to brain health further on in life^7^. On average, a collegiate football player receives about 1,000 subconcussive head impacts in a single season ^8^, and it is estimated that only 1 in 9 symptom-provoking concussions are reported ^9,10^. Despite the high occurrence rates, we still lack a proper understanding of how to reverse the detrimental consequences of repetitive mTBI. For example, it was described that athletes heavily exposed to repetitive mTBI exhibit impaired cognitive function and neuropathological consequences later in life^11-13^.

Interestingly, the current evidence summarized in a review and meta-analysis refers to professional/elite players only and points towards an association between sustaining a concussion and inferior cognitive function in rugby, American football, and boxing. However, the data included refers to professional/elite players only, and it is unclear to what extent this is relevant in the clinic^14^. Therefore, high-quality studies are urgently needed to assess further the association between sustaining a concussion and cognitive impairment later in life.

The brain and peripheral inflammation triggered after mTBI plays an essential role in the short and long-term effects of the trauma^15^. Many inflammatory biomarkers are substantially altered after concussion when compared with non-injured athletes^16^. Concussions are associated with an increase in systemic inflammatory molecules ^17^. As such, a promising approach for brain recovery is to understand better the mechanisms through which the inflammation generated in the periphery after head injury leads to the development of short- and long-term cognitive deficits. Therefore, developing reliable biomarkers for concussions and repetitive subconcussive hits is critical to improving diagnosis and constitutes the first step to investigate new anti-neuroinflammatory therapeutic approaches.

Standard neuroimaging studies in injured patients typically do not show short- or long-term brain damage after a concussion. However, abnormal TBI patterns of magnetic resonance imaging (MRI) activation have been observed in some studies of sports-related concussions^18^. Several advanced imaging studies have been evaluated in the past couple of decades to study a functional or pathophysiologic change rather than a structural one ^19^. More sophisticated neuroimaging techniques such as diffusion tensor imaging (DTI) have detected changes in white matter tracts in individuals with a history of a SRC that is often widespread^20,21^. There are several limitations to identifying blood biomarkers since proteins released by the brain are only detectable at low plasma concentrations. They undergo proteolytic degradation in the blood, and their levels are affected by elimination through the liver or kidney^22^. However, it is necessary to identify reliable brain injury biomarkers^23^, which reflect the severity of the concussion to determine the diagnosis or prognosis in collision sports athletes.

Head trauma leads to both the acute and chronic disruption of the intestinal barrier and the subsequent appearance of bacterial endotoxins in the blood^24^. The gastrointestinal and metabolic disruptions that often accompany brain trauma can be explained by the bidirectional link between the central nervous system (CNS) and the enteric nervous system (e.g., the brain-gut axis). Patients with moderate to severe brain injury often show neurogenic gut dysfunction due to a lack of CNS control over the gastrointestinal tract^25^, suggesting a possible involvement of the gut microbiota in head injury outcomes. The gut microbiota is an essential neuromodulator of brain-gut signaling and can affect brain inflammation resulting from brain injury^26^.

Severe TBI patients treated with *Lactobacillus*-rich probiotic supplementation have decreased gastrointestinal dysfunction and a shortened time spent in intensive care^27^. These observed benefits are commonly attributed to probiotic-induced reductions in systemic and central inflammation^28^. Similarly, thinking about a post-concussion clinical treatment, novel preventive and therapeutic strategies, including nutritional diets, microbiota manipulations with probiotics or prebiotics, or strengthening the enteric barrier could be applied to modulate the intestinal microbiota and ultimately improve both cognitive and functional TBI health outcomes. In addition, changes in the gut microbiota (or dysbiosis) have previously been correlated with developing certain diseases, including neuropsychiatric disease^29^. Dysbiosis of the intestinal microbiota has been described in patients with chronic severe TBI^30,31^ and related to alterations in the permeability of the blood-brain barrier and microglia activation^32^, as well as changes in the microbiome composition^33^. Although gut dysbiosis after a single, severe TBI has been documented, it has not been studied in SRC that are almost exclusively mild TBIs or in subconcussive hits in athletes exposed to repetitive head impacts. Therefore, we suggest that a single diagnosed concussion or multiple subconcussive hits sustained during a collegiate football season may disrupt the gut microbiome.

We utilize computational and statistical approaches to identify several changes in gut microbiomes in athletes after a concussion, or during collision sports activity, and investigate their relationship to inflammatory blood biomarkers. By assessing collegiate athletes longitudinally, we aimed to identify changes to the microbiota that are relevant pathophysiology of head trauma and can potentially provide a better tool to predict clinical outcomes.

## RESULTS

### Demographic and clinical characteristics

This study included 33 male Division I collegiate football players, four of whom suffered from a diagnosed concussion while enrolled in the study. Details of the study population are provided in Table 1 and the Methods section. Three data collection timepoints were established: two during the athletic activities (mid- and post-season) and an off-season collection 86 days after the last game of the season (Fig. 1). Additional data collection was performed for the concussed players within 48 hours after diagnosis.

**Table 1.**
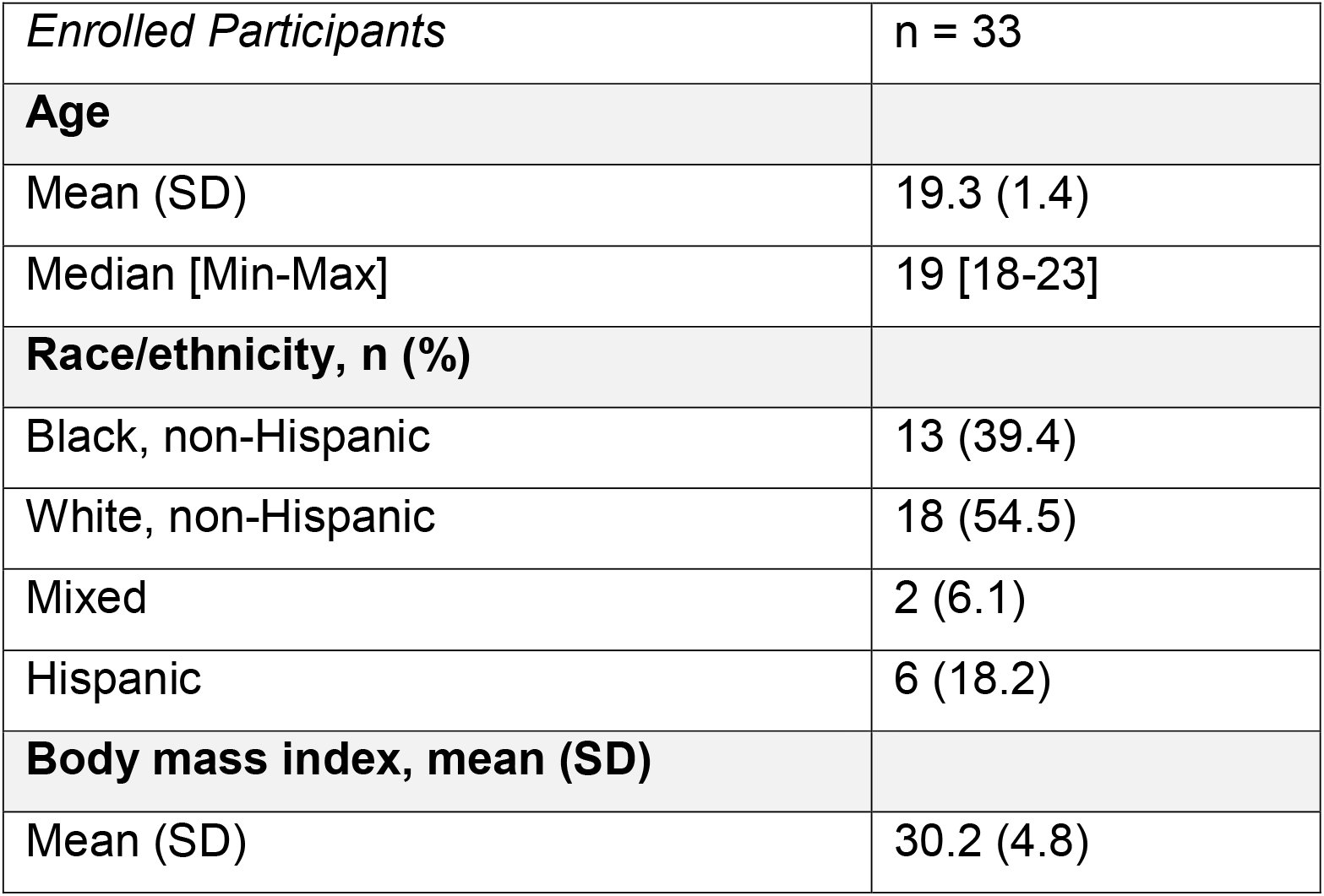
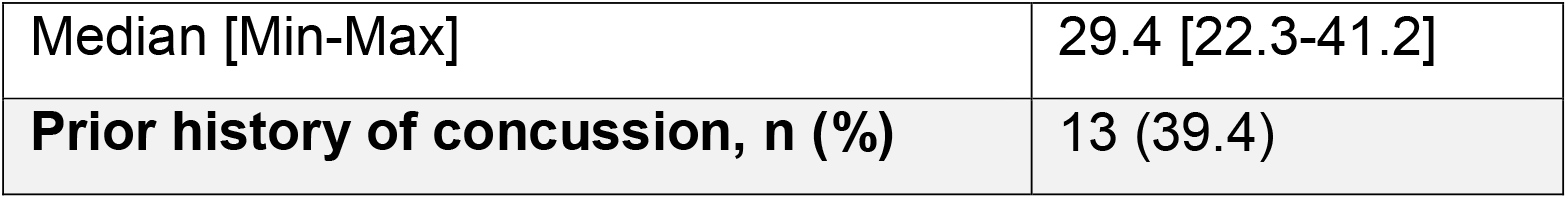
Demographic data in the enrolled participant groups.

|  |  |
| --- | --- |
| *Enrolled Participants* | n = 33 |
| **Age** |  |
| Mean (SD) | 19.3 (1.4) |
| Median [Min-Max] | 19 [18-23] |
| **Race/ethnicity, n (%)** |  |
| Black, non-Hispanic | 13 (39.4) |
| White, non-Hispanic | 18 (54.5) |
| Mixed | 2 (6.1) |
| Hispanic | 6 (18.2) |
| **Body mass index, mean (SD)** |  |
| Mean (SD) | 30.2 (4.8) |
| Median [Min-Max] | 29.4 [22.3-41.2] |
| **Prior history of concussion, n (%)** | 13 (39.4) |

**Figure 1.**
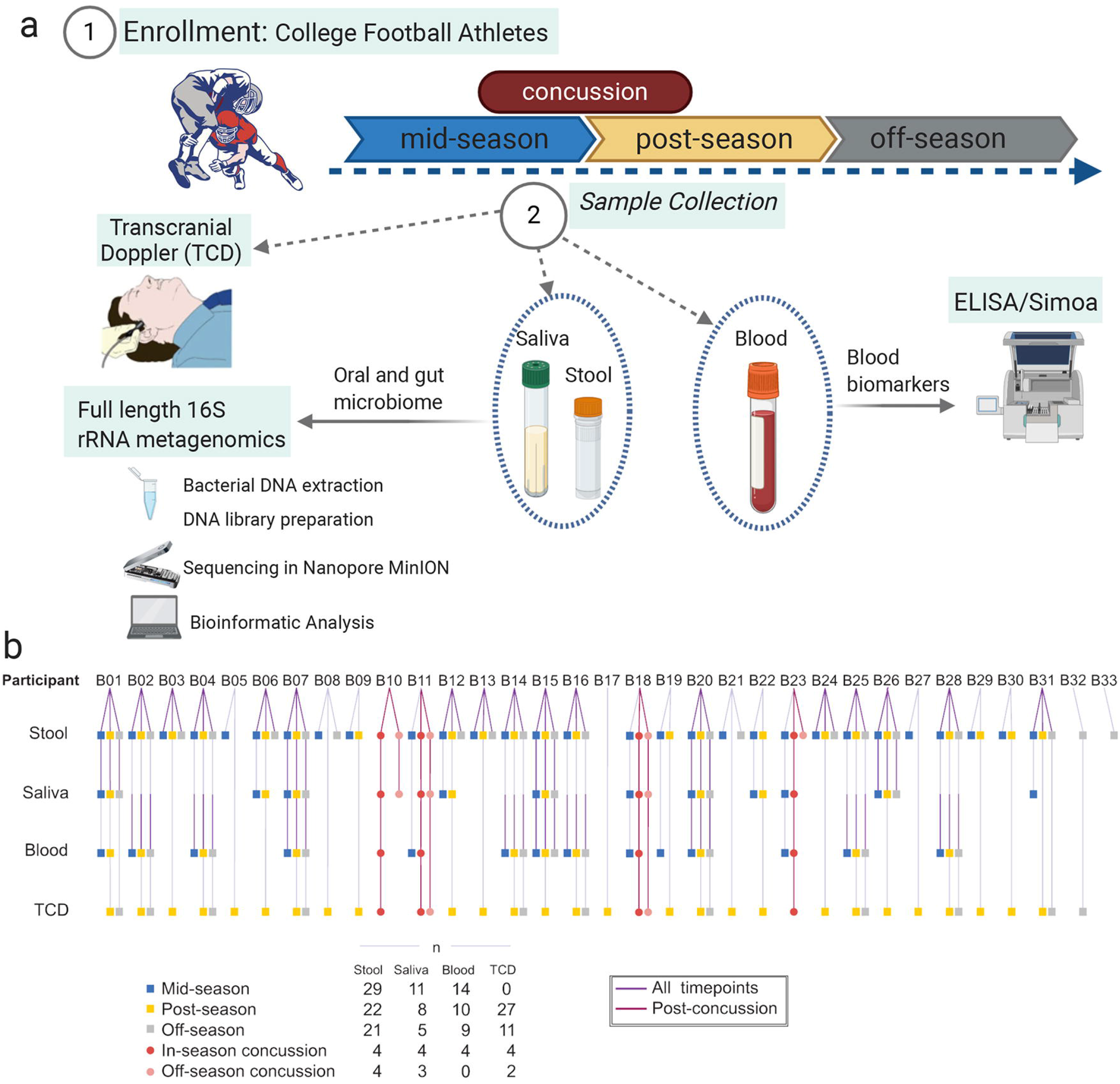
Study Design and participants. (a) Saliva, stool, and blood samples were collected from collegiate football players athletes (n=33) during the football season. Transcranial doppler imaging measurements were performed and protein analysis was completed using a commercial ELISA and single molecule array. Bacterial DNA was extracted and 16S rRNA sequencing analysis was performed using the Nanopore platform. (b) Breakdown of samples received from each of the participants in the study at each of the designated timepoints.

### Changes in the human gut microbiome and corresponding pathways across the football season

Microbial DNA was extracted from fecal samples, and next-generation 16S rRNA gene sequencing was performed using the Nanopore MinION platform, which has been shown to provide a more accurate taxonomic classification at the species level than Illumina MiSeq^34^. Long-read 16S rRNA amplicons covering the V1-V9 hypervariable region generated reads of approximately 1500 bp. Sequences were then trimmed using Porechop and classified with Kraken ^35^, which yielded 23,249,965 classified reads and led to 1,021 species being identified after quality filtering. For the longitudinal analysis of gut microbiota changes across the football season, we excluded any samples collected following a concussion and subsequently grouped all remaining samples according to the three collection timepoints (mid-season, n=29; post-season, n=22, and off-season, n=21). Then, we investigated the differences in gut microbiota community structure across timepoints by calculating beta and alpha diversity metrics. Plotting of weighted UniFrac distances by principal coordinate analysis (PCoA) revealed overlap between the three timepoints, with no statistically significant differences evaluated by analysis of similarity (ANOSIM) (R=0.0064, p=0.3622; Fig. 2a). Further, the alpha diversity at the species level based on the Simpson and Shannon metrics did not significantly differ between the time points (p=0.178 and p=0.425, respectively) (Fig. 2b). Next, we evaluated the gut microbiota composition at the phylum, family, and genus level for all three-time points. Overall, the relative abundance of prevalent taxa was similar between the mid- and post-seasons. However, some shifts were observed in the off-season compared to the other timepoints (Fig. 2c). A random-effects multivariate analysis that only included data from non-concussed athletes who had provided a sample at all three timepoints (n=17) was performed using MaAsLin2. Mid- to post-season and post- to off-season were compared in two independent analyses (Supplemental Table 1). This model identified 8 bacteria species with a statistically significant change in abundance between the post- and off-season timepoints (i.e., having an FDR-corrected p-value <0.05). Supplemental Table 1 shows these species but also extends to show all species with an FRD-corrected p-value as high as 0.125. It is noteworthy that of the 17 species shown, all of them had decreased in relative abundance during the off-season (Fig. 2d). Microbial taxa at the family, genus, and species level meeting both statistical significance criteria and expressing prevalent abundance (mean > 1% in at least one group at family or genus level, >0.1% at species level) are represented in Fig 2e. Of note, the species *Anthrobacter sp*. YN *and Desulfirispirillum indicum* decrease in relative abundance during the off-season compared to the post-season (q=0.027 and 0.039, respectively).

**Figure 2.**
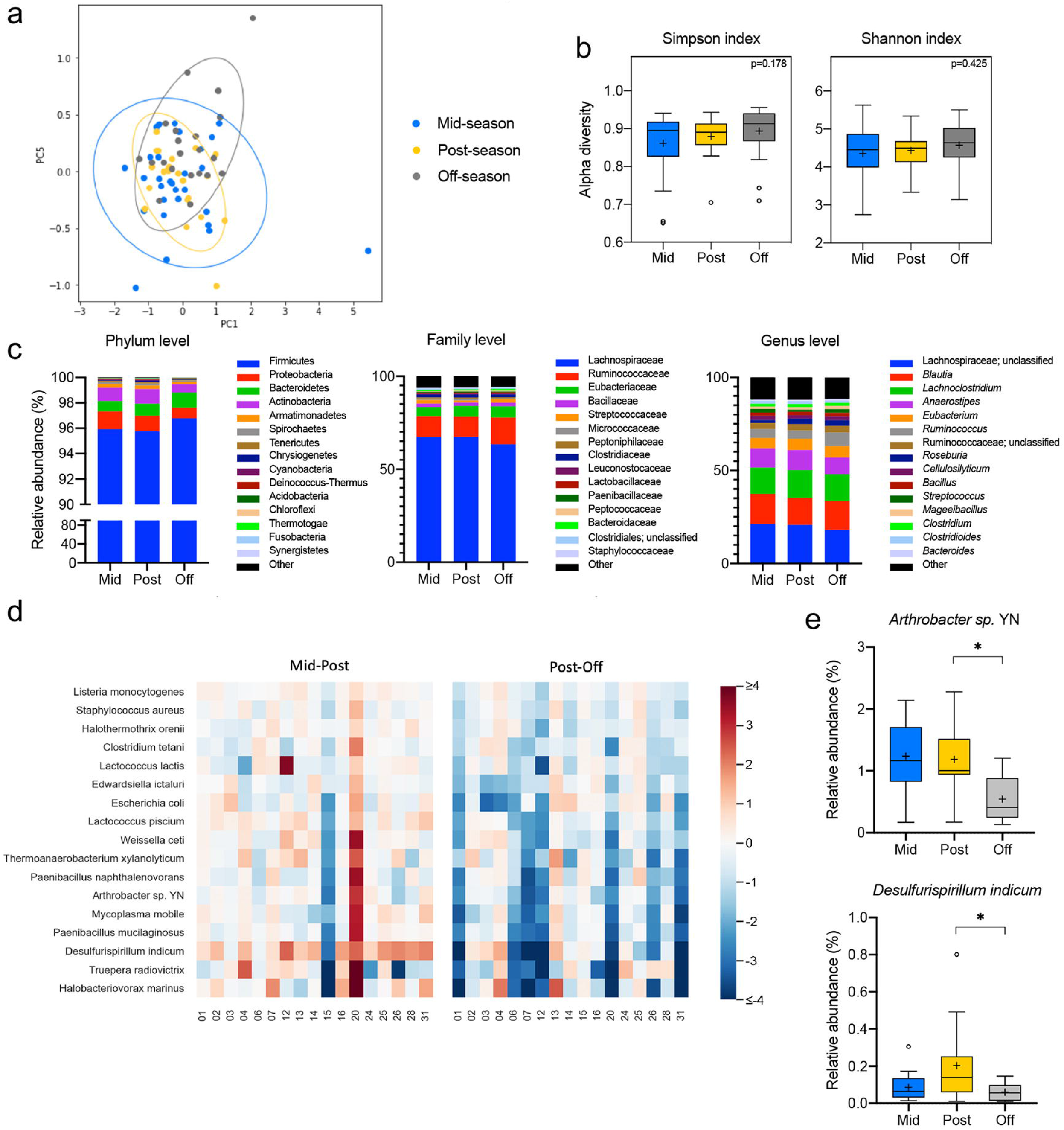
Diversity and taxonomic analysis along the sports season in non-concussed football athletes. (a) Plots of Beta diversity analysis of the fecal microbiota from athletes who did not experience a diagnosed concussion during the football season. The Principal Coordinate Analysis (PCoA) ordination plot was based on Weighted UniFrac distances for the mid-(n=29, blue), post-(n=22, yellow), and off-season (n=21, gray) groups. Confidence ellipses represent the 95% confidence interval for each timepoint. Group dissimilarities were evaluated by Analysis of Similarity (ANOSIM). (b) Shannon and Simpson alpha diversity indices at the species level for the mid-(n=29, blue), post-(n=22, yellow), and off-season (n=21, gray) timepoints. In the box and whisker plots, the cross represents the mean. Significance was determined by Kruskal-Wallis followed by Dunn’s multiple comparisons test. (c) Relative abundances of the top 15 phyla, families, and genera in the fecal microbiota of the non-concussed athletes through the mid-(n=29, blue), post-(n=22, yellow), and off-(n=21, gray) seasons. (d) Heatmap of the log2 fold changes for the species that are significantly altered across the sport season in the non-concussed athletes. Statistical testing by random-effects multivariate analysis included the samples from athletes who participated in all collection timepoints (n=17). All statistically significant changes were identified between the post- and off-season. (e) Relative abundances at the mid-(n=17, blue), post-(n=17, yellow), and off-season (n=17, gray) timepoints of the prevalent taxa (abundance >1% in at least one of the groups at the family and genus level, >0.1% at the species level) showing significant alterations in the random-effects multivariate analysis. In the box and whisker plots, the cross represents the mean. *q<0.05; ***q<0.001.

We next inferred the functional composition of the gut microbiota based on the 16S data using PICRUSt2^36^. Changes in abundance of MetaCyc pathways throughout the football season in non-concussed football athletes (n=17) were identified by two independent MaAsLin2 random-effects multivariate analyses comparing the mid- to post-season and post- to the off-season. Results revealed 251 pathways that were differentially abundant between groups, with all significant changes (q<0.05) occurring between the post- and off-season (Supplemental Table 3). Identification of the parent classes for the 251 differentially abundant pathways revealed that they are predominantly related to aromatic compound degradation, amino acid biosynthesis and degradation, fatty acid biosynthesis, and biosynthesis of electron carriers, such as quinol and quinone. In particular, the pathways with the highest fold changes between the post- and off-season are most notably involved in the degradation of sugars and aromatic compounds (Fig. 3).

**Figure 3.**
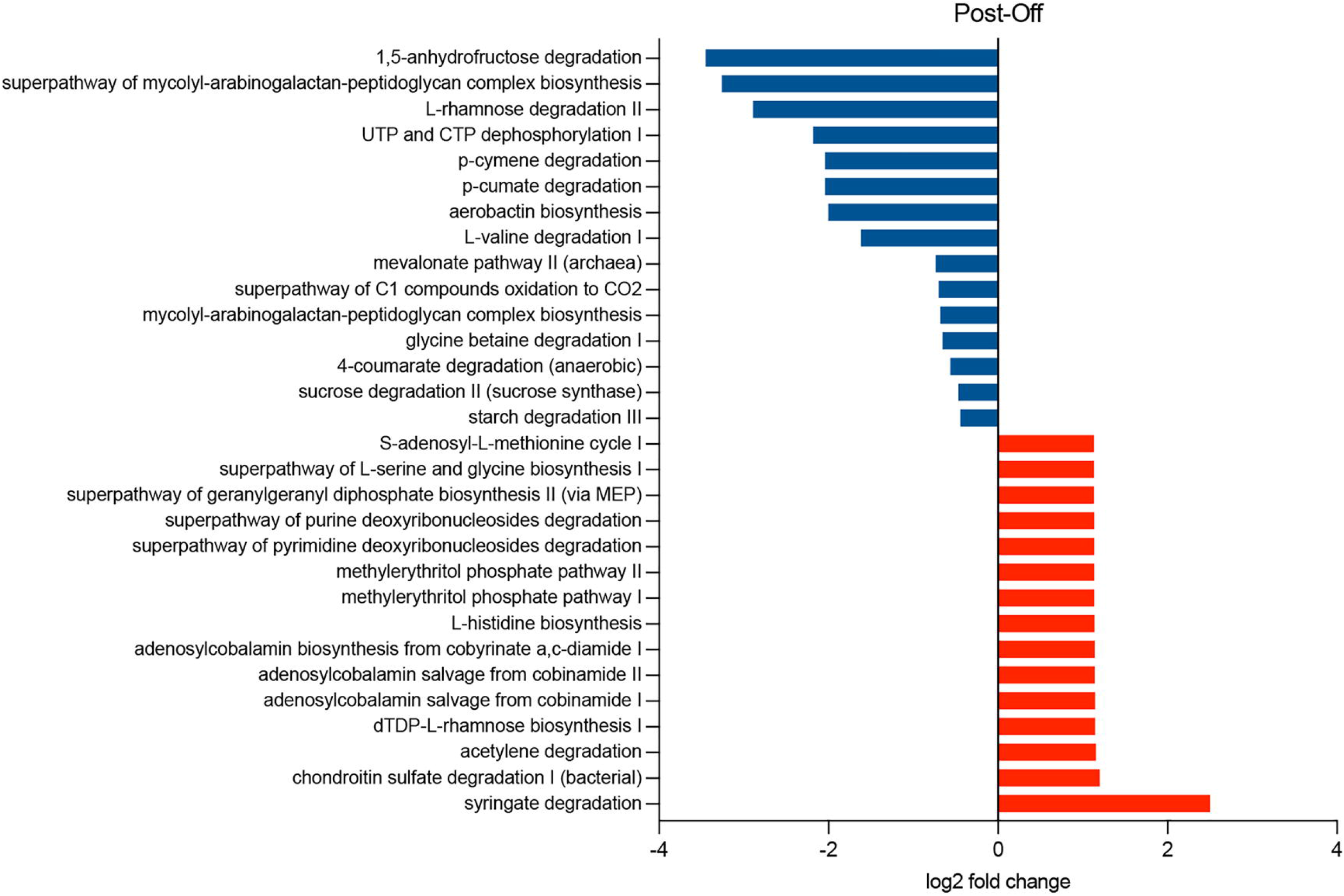
Pathway abundance prediction by PICRUSt2 of the fecal microbiota throughout the football season in non-concussed athletes. Log2 fold change of the abundances of the MetaCyc pathways showing significant differences (q<0.05) by random-effects multivariate analysis of the samples from athletes who provided samples at all three timepoints (n=17). All statistically significant changes were identified between the post- and off-season. Top 15 log2 fold changes towards increase (red) and reduction (blue) are represented in the bar plot.

### Relationship between the gut microbiota and a single concussion

We next evaluated the changes in the gut microbiome associated with concussion. Since mid-and post-season samples proved similar in the longitudinal analysis, both timepoints were combined to form an “in-season” group. Samples were then divided into four groups: in-season (n=51), in-season concussion (n=4), off-season (n=21), and off-season concussion (n=4) (Fig. 1b). The in-season concussion group consists of the samples from the four athletes who suffered a concussion collected within 24 to 48 hours following the diagnosed concussion. In contrast, the off-season concussion group includes the samples from the same four athletes who received a concussion collected in the later off-season timepoint.

Beta diversity analysis showed significant differences in the gut microbiota structure between groups (R=0.2309, p<0.001). Pairwise ANOSIM analysis indicated that samples from concussed and non-concussed players were significantly different for both in-(R=0.3959, p=0.0127) and off-season (R=0.5711, p=0.0024) comparisons (Fig 4a). Additionally, greater alpha diversity was observed in the concussed athletes when compared to their non-concussed teammates, for both the in-season (Simpson, p=0.0082; Shannon p=0.0043) and off-season (Simpson, p=0.0435; Shannon p=0.0329) comparison (Fig. 4b). The abundance of the most prevalent genera and families from the gut microbiota was similar within each group of participants (concussed and non-concussed athletes) between the in- and the off-season timepoints (Fig. 4c). At the phylum level, fecal samples collected post-concussion during the in-season exhibited an overall distinct distribution of the most abundant phyla compared to the rest of the groups represented (Fig. 4c).

**Figure 4.**
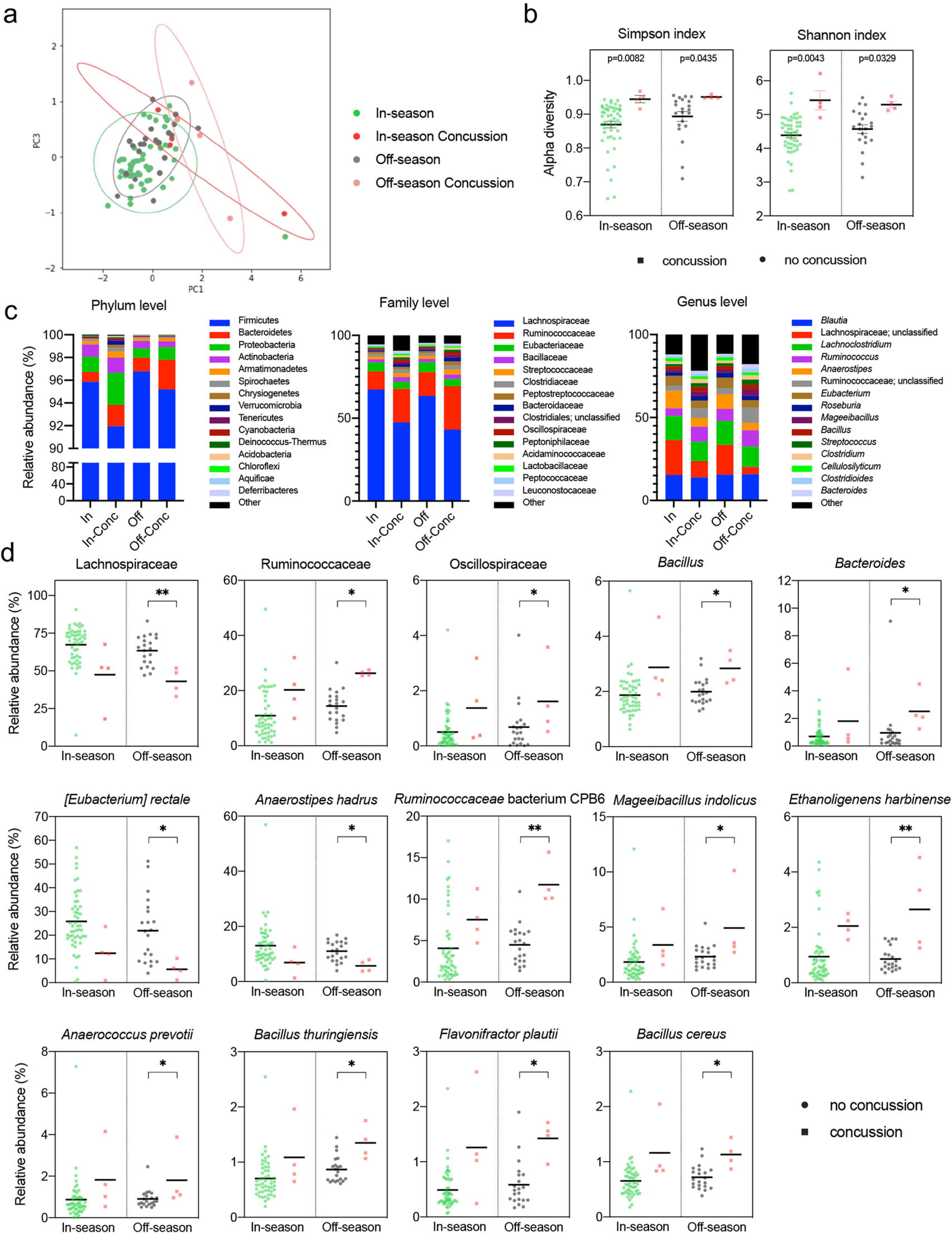
Diversity and taxonomic analysis in athletes after a single concussion. (a) Beta diversity analysis of fecal microbiota from athletes who experienced a diagnosed concussion during the football season. Principal Coordinate Analysis (PCoA) ordination plot was based on Weighted UniFrac distances for the in-season (n=51, green), in-season concussion (n=4, red), off-season (n=21, gray), and off-season concussion (n=4, pink) groups. Confidence ellipses represent the 95% confidence interval for each timepoint. Group dissimilarities were evaluated by Analysis of Similarity (ANOSIM). (b) Shannon and Simpson alpha diversity indices at the species level for concussed (n=4 in-season, red and off-season, pink) and non-concussed (n=51 in-season, green and n=21 off-season, gray) football athletes. In the box and whisker plots, the cross represents the mean. Significance was determined by Kruskal-Wallis followed by Dunn’s multiple comparisons test. (c) Relative abundances of the top 15 phyla, families, and genera in the fecal microbiota for the in-season (n=51, green), in-season concussion (n=4, red), off-season (n=21, gray) and off-season concussion (n=4, pink) groups. (d) Relative abundances of the prevalent taxa (abundance >1% in at least one of the groups at the family and genus level, >0.1% at the species level) showing significant alterations in the random-effects multivariate analysis for concussed (n=4 in-season, red and off-season, pink) and non-concussed (n=51 in-season, green and n=21 off-season, gray) athletes. In the box and whisker plots, the cross represents the mean. *q<0.05; **q<0.01.

A fixed-effects model in MaAsLin2 was used to identify specific microbial taxa with statistically significant differences in abundance between the concussed and non-concussed players at the in- and off-season timepoints (Supplemental Table 2). The model identified 9 species with a significant difference in the off-season model, while none showed significance in the in-season analysis. The relative abundance of the family *Lachnospiraceae* was decreased in the concussed athletes compared to their non-concussed teammates (q=0.003), whereas the families *Ruminococcaceae* (q=0.028) and *Oscillospiraceae* (q=0.028) were increased (Fig. 4d). At the genus level, Bacillus and Bacteroidetes were increased in the concussion group for the off-season analysis (q=0.025 and q=0.023, respectively). The species *Eubacterium rectale and Anaerostipes hadrus*, both belonging to the *Lachnospiraceae* family, significantly reduced relative abundance in concussed athletes (q=0.014 and q=0.034, respectively). On the contrary, seven bacterial species, including *Ruminococcaceae bacterium* CPB6 (q=0.007), *Mageeibacillus indolicus* (q=0.021), *Ethanoligenes harbinense* (q=0.007), *Anaerococcus prevotii* (q=0.032), *Bacillus thuringiensis* (q=0.014), *Flavonifractor plautii* (q=0.014) and *Bacillus cereus* (q=0.017), showed a greater relative abundance in the concussion group.

To investigate the functional changes in the gut microbiota following concussion, we examined pathway abundances generated with PICRUSt2. MaAsLin2 multivariate analysis between the concussed and non-concussed groups identified two and eight differentially abundant pathways in the in-season and off-season, respectively (Fig. 5, Supplemental Table 3). These results indicate that the major changes between off-season concussion and non-concussion included: acetylene degradation, N10-formyl-tetrahydrofolate biosynthesis, NAD salvage pathway, thiamin salvage, flavin biosynthesis, D-galacturonate degradation, beta D-glucuronide and De-glucoronate degradation, hexuronide and hexuronate degradation, and adenosine nucleotides degradation. On the other hand, the major changes between the in-season concussion and in-season included: coumarins biosynthesis and 1, 3-propanediol biosynthesis (Fig. 5). Notably, among the pathways altered in the off-season, sugar acid degradation pathways had significantly reduced relative abundances in the concussion group compared to the non-concussed athletes.

**Figure 5.**
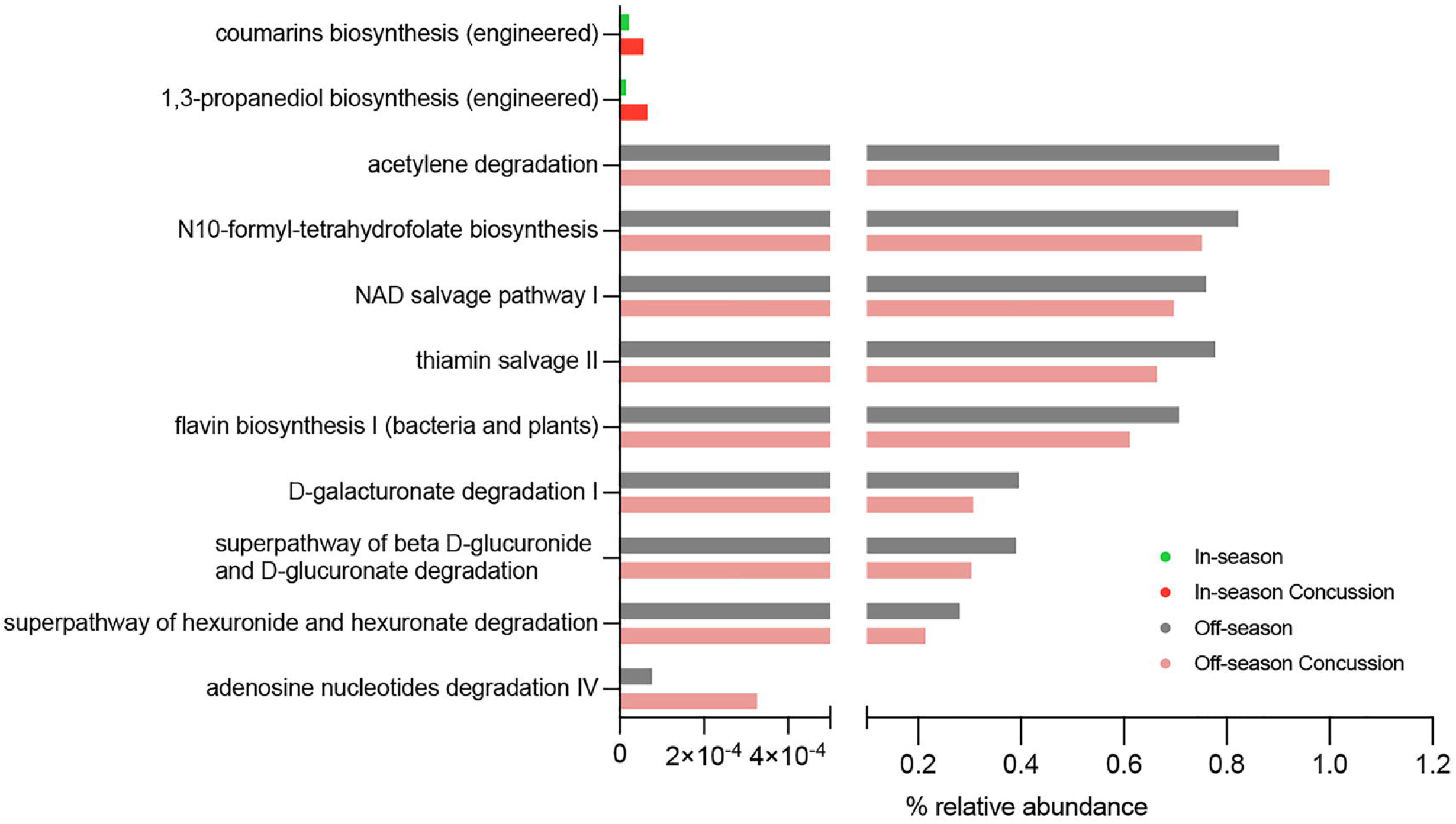
Pathway abundance prediction by PICRUSt2 of the fecal microbiota following concussion. Abundance of the MetaCyc pathways showing significant differences (q<0.05) by random-effects multivariate analysis in the in-season (n=51, green), in-season concussion (n=4, red), off-season (n=21, gray), and off-season concussion (n=4, pink) groups.

### No alterations in the oral microbiota during the longitudinal study or after a concussion

To study the changes in the oral microbiome taking place as the football season progressed, we analyzed 16S rRNA gene sequencing data from the saliva of non-concussed players (mid-season, n=11; post-season, n=8; and off-season, n=5) (Fig. 1b). Nanopore MinION long-read 16S rRNA sequencing yielded 6,967,795 classified reads with 793 species identified after quality filtering. The structure of the saliva microbial community was similar between groups, as indicated by the beta diversity analysis (R=-0.0178, p=0.5415) (Fig. 6a). Similarly, no significant differences in alpha diversity were observed across the three timepoints analyzed (Simpson, p=0.779; Shannon, p=0.975) (Fig. 6b). The overall shifts in the oral microbiota composition at the phylum, family, and genus level do not appear to be consistent across the three timepoints analyzed (Fig. 6c).

**Figure 6.**
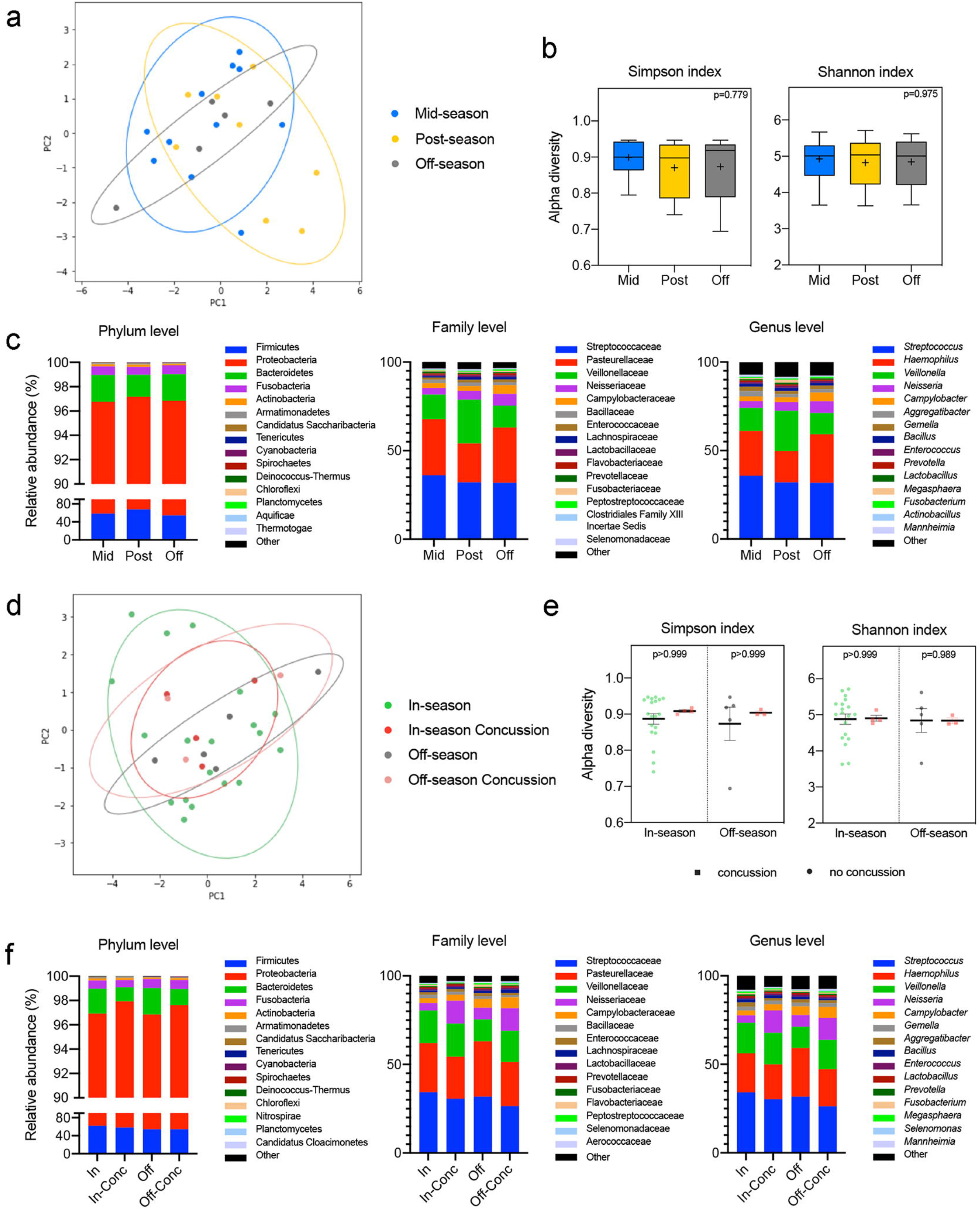
Diversity and taxonomic analysis of salivary microbiome throughout the season in football athletes. (a) Beta diversity analysis of the saliva microbiota from athletes who did not experience a diagnosed concussion during the football season. Principal Coordinate Analysis (PCoA) ordination plot was based on Weighted UniFrac distances for the mid-(n=11, blue), post-(n=8, yellow), and off-season (n=5, gray) groups. Confidence ellipses represent the 95% confidence interval for each timepoint. Group dissimilarities were evaluated by Analysis of Similarity (ANOSIM). (b) Shannon and Simpson alpha diversity indices at the species level for the mid-(n=11, blue), post-(n=8, yellow), and off-season (n=5, gray) timepoints. In the box and whisker plots, the cross represents the mean. Significance was determined by Kruskal-Wallis followed by Dunn’s multiple comparisons test. (c) Relative abundances of the top 15 phyla, families, and genera in the oral microbiota of the non-concussed athletes through the mid-(n=11, blue), post-(n=8, yellow), and off-(n=5, gray) seasons. (d) Beta diversity analysis of the saliva microbiota from athletes who experienced a diagnosed concussion during the sports season. Principal Coordinate Analysis (PCoA) ordination plot was based on Weighted UniFrac distances for the in-season (n=19, green), in-season concussion (n=4, red), off-season (n=5, gray), and off-season concussion (n=3, pink) groups. Confidence ellipses represent the 95% confidence interval for each timepoint. Group dissimilarities were evaluated by Analysis of Similarity (ANOSIM). (e) Shannon and Simpson alpha diversity indices at the species level for the concussed (n=4 in-season, red and n=3 off-season, pink) and non-concussed (n=19 in-season, green and n=5 off-season, gray) football athletes. In the box and whisker plots, the cross represents the mean. Significance was determined by Kruskal-Wallis followed by Dunn’s multiple comparisons test. (f) Relative abundances of the top 15 phyla, families and genera in the oral microbiota for the in-season (n=19, green), in-season concussion (n=4, red), off-season (n=5, gray), and off-season concussion (n=3, pink) groups.

We next assessed whether the oral microbiome was altered in concussed athletes (in-season, n=4; off-season, n=3) compared to their non-concussed teammates (in-season, n=19; off-season n=5) (Fig. 1b). No significant differences were found in either beta diversity (R=-0.0464; p=0.6387; Fig. 6d, e) or alpha diversity (Simpson, p>0.999; Shannon p>0.999 in-season, p=0.989 off-season). At the phylum, family, and genus level, the most abundant taxa are overall comparable in abundance between the in- and the off-season within the same group of athletes (concussed and non-concussed) (Fig. 6f). In conclusion, saliva microbiota biomarkers remained close to their baseline values through all timepoints and after a concussion.

### No changes in the optic nerve sheath diameter or cerebral blood flow measurements

There were no significant differences in the optic nerve sheath diameters (ONSD) or transcranial Doppler (TCD) parameters in the right (R) and left (L) cerebral blood flow velocities (CBFV) (velocity without breath-holding (BH), the velocity with BH, holding time, anterior cerebral artery (ACA), posterior cerebral artery (PCA), internal carotid artery (ICA), L and R PI without BH, and with BH, and diameters of the optic nerve sheath (ONSD) for R and L (diameter and BHI). A single sonographer assessed TCD measurements at the post-season and off-season timepoints (Table 2). No clinical concussion participants underwent clinical TCD testing, and no meaningful differences in velocity or diameter were observed across the timepoints.

**Table 2.**
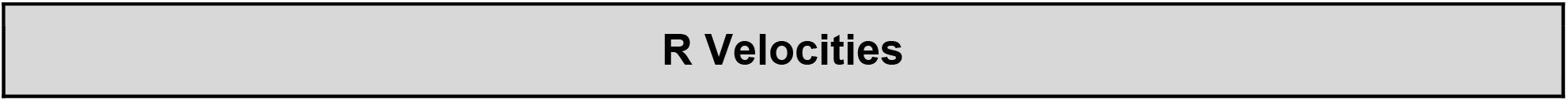
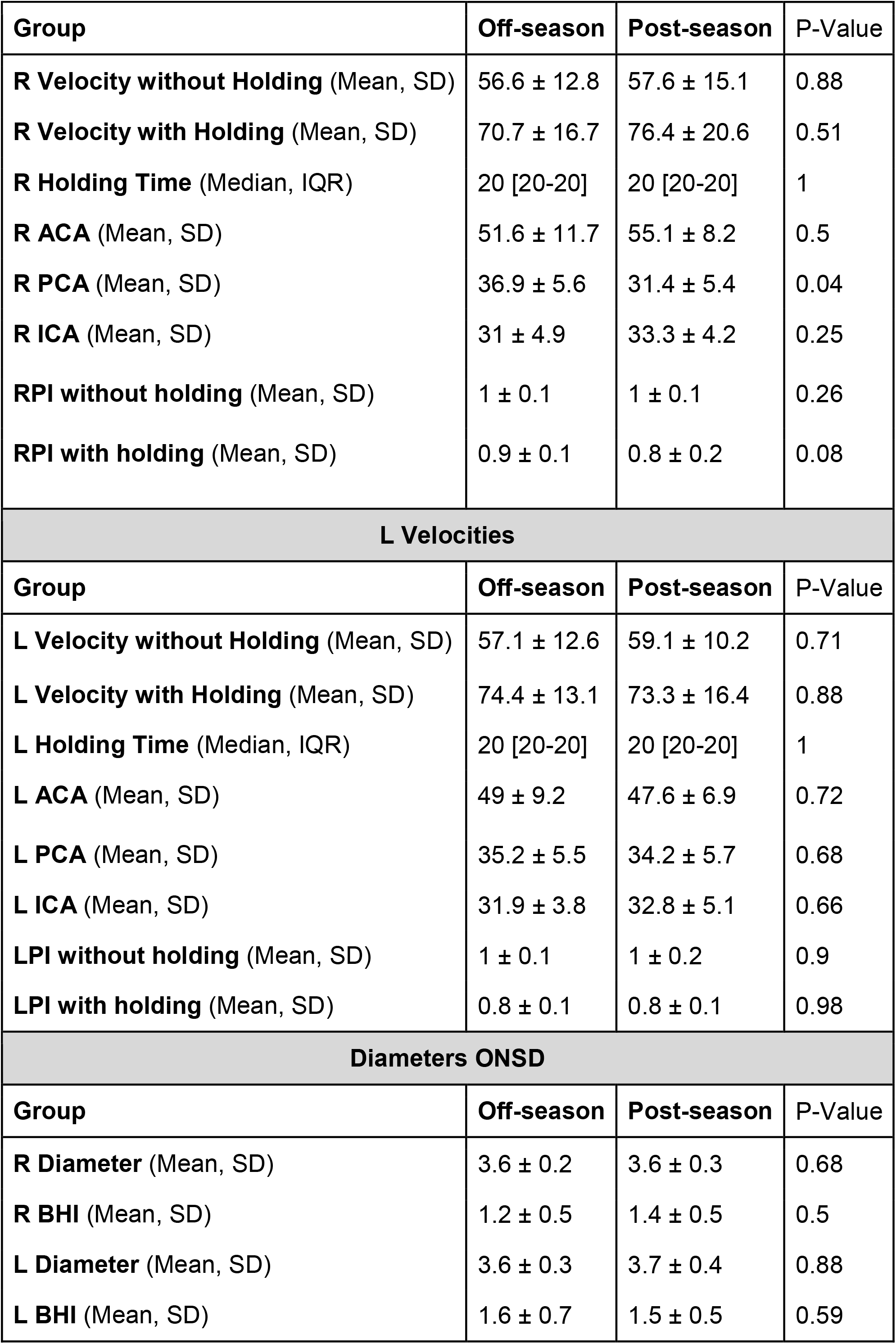
Transcranial doppler imaging optic nerve sheath diameter statistics. Right (R) and left (L), breath-holding (BH), anterior cerebral artery (ACA), posterior cerebral artery (PCA), internal carotid artery (ICA), optic nerve sheath (ONSD).

| Group | Off-season | Post-season | P-Value |
| --- | --- | --- | --- |
| **R Velocity without Holding** (Mean, SD) | 56.6 ± 12.8 | 57.6 ± 15.1 | 0.88 |
| **R Velocity with Holding** (Mean, SD) | 70.7 ± 16.7 | 76.4 ± 20.6 | 0.51 |
| **R Holding Time** (Median, IQR) | 20 [20-20] | 20 [20-20] | 1 |
| **R ACA** (Mean, SD) | 51.6 ± 11.7 | 55.1 ± 8.2 | 0.5 |
| **R PCA** (Mean, SD) | 36.9 ± 5.6 | 31.4 ± 5.4 | 0.04 |
| **R ICA** (Mean, SD) | 31 ± 4.9 | 33.3 ± 4.2 | 0.25 |
| **RPI without holding** (Mean, SD) | 1 ± 0.1 | 1 ± 0.1 | 0.26 |
| **RPI with holding** (Mean, SD) | 0.9 ± 0.1 | 0.8 ± 0.2 | 0.08 |
| **L Velocities** |  |  |  |
| Group | Off-season | Post-season | P-Value |
| **L Velocity without Holding** (Mean, SD) | 57.1 ± 12.6 | 59.1 ± 10.2 | 0.71 |
| **L Velocity with Holding** (Mean, SD) | 74.4 ± 13.1 | 73.3 ± 16.4 | 0.88 |
| **L Holding Time** (Median, IQR) | 20 [20-20] | 20 [20-20] | 1 |
| **L ACA** (Mean, SD) | 49 ± 9.2 | 47.6 ± 6.9 | 0.72 |
| **L PCA** (Mean, SD) | 35.2 ± 5.5 | 34.2 ± 5.7 | 0.68 |
| **L ICA** (Mean, SD) | 31.9 ± 3.8 | 32.8 ± 5.1 | 0.66 |
| **LPI without holding** (Mean, SD) | 1 ± 0.1 | 1 ± 0.2 | 0.9 |
| **LPI with holding** (Mean, SD) | 0.8 ± 0.1 | 0.8 ± 0.1 | 0.98 |
| **Diameters ONSD** |  |  |  |
| Group | Off-season | Post-season | P-Value |
| **R Diameter** (Mean, SD) | 3.6 ± 0.2 | 3.6 ± 0.3 | 0.68 |
| **R BHI** (Mean, SD) | 1.2 ± 0.5 | 1.4 ± 0.5 | 0.5 |
| **L Diameter** (Mean, SD) | 3.6 ± 0.3 | 3.7 ± 0.4 | 0.88 |
| **L BHI** (Mean, SD) | 1.6 ± 0.7 | 1.5 ± 0.5 | 0.59 |

### Changes in blood biomarkers across the football season

Eleven participants provided blood samples at each of the three timepoints, enabling us to perform a longitudinal analysis of blood serum biomarkers. Specifically, repeated measures ANOVAs were used to determine if the levels of several biomarkers in serum changed across the football season. Results revealed no significant differences for S100β (F(2, 16)=1.087, p=0.361), serum amyloid A (SAA) (F(2, 16)=1.433, p=0.268) and neurofilament light chain (NF-L) (F(2,16)=1.262, p=0.310) (Fig. 7). However, serum glial fibrillary acidic protein (GFAP) was significantly decreased in both the mid-season and the post-season compared to the off-season (p=0.00813 and p=0.0437, respectively; F(2,16)=6.597, p=0.0081) (Fig. 7c). We also compared the biomarker levels in serum in the four samples collected in-season after a concussion to the samples from the rest of the athletes. There was no statistical difference between the groups tested for S100β, SAA, glial fibrillary acidic protein (GFAP), and neurofilament light (NF-L) (p=0.312, p=0.496, p=0.431, and p=0.616, respectively) (Fig 7e-h). However, we did detect a significant correlation between the concentration of the serum biomarkers S100β and SAA in the longitudinal samples of non-concussed athletes with the abundance of the bacterium species *Eubacterium rectale* (Fig. 7i).

**Figure 7.**
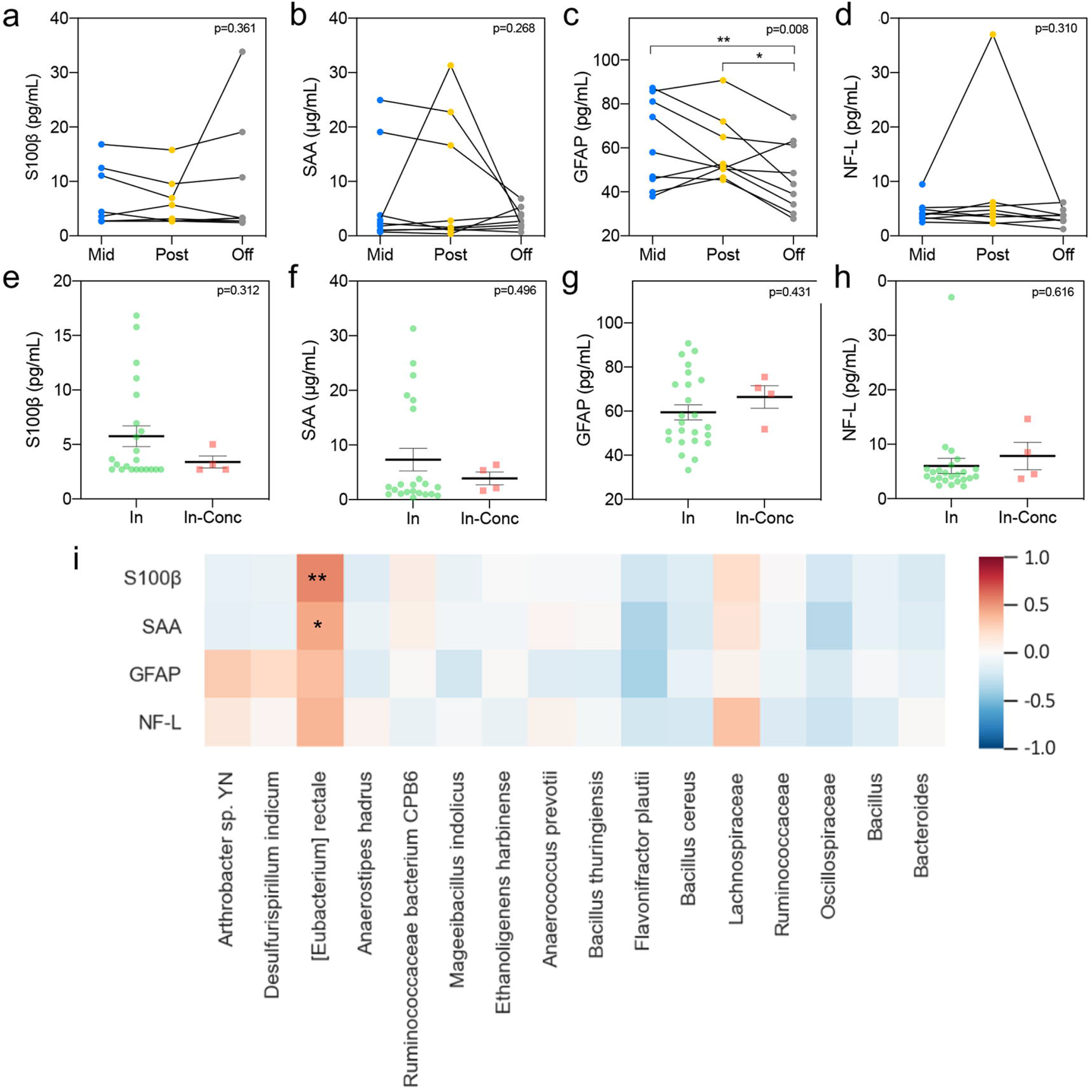
Serum levels of biomarkers related to brain injury. (a-d) S100β (a), SAA (b), GFAP (c), and NF-L (d) serum concentrations represented longitudinally for each of the football athletes who did not experience a diagnosed concussion, at mid-(n=9, blue), post-(n=9, yellow), and off-season (n=9, gray). Significance was determined by Repeated Measures ANOVA followed by Tukey multiple comparisons test (*p<0.05). (e-h) S100β (e), SAA (f), GFAP (g), and NF-L (h) serum concentrations after diagnosed concussion (n=4, red) compared to the values in the in-season (n=24, green) for the rest of the players in the study. Significance was determined by an unpaired T-test. SAA: Serum amyloid A; GFAP: Glial fibrillary acidic protein; NF-L: Neurofilament light. (i) Heatmap of the Pearson’s correlation coefficients between the concentration of serum biomarkers throughout the sports season and the relative abundance of species and genus significantly changed in the gut microbiome analysis. *p<0.05; **p<0.01.

## DISCUSSION

The results of our study identify alterations to the gut microbiome profiles in collegiate student-athletes who have suffered a single concussion or were subject to repeated subconcussive blows to the head during the active period of a football season. Currently, collegiate athletes are removed from play if signs and symptoms of traumatic head or cervical injury or concussion are observed by the sideline medical staff. However, since an asymptomatic player may still have a mild brain injury, indicative biomarkers corroborating a brain injury diagnosis are needed. This study identifies specific bacteria that decrease in abundance after concussion (*Eubacterium rectale and Aerostipes hadrus)*, suggesting the gut microbiome as a potentially useful biomarker for concussions. Even subtle changes in gut microbiota, and the metabolites that are produced, may affect gut health, athletic performance^37^ and exercise intensity^38^. There is also a clear need for novel diagnostics to implement suitable recovery periods and ultimately reduce the risks of prolonged overexposure and inflammation due to concussion and subconcussive blows to the head.

Changes in the gut microbiota in response to the inflammatory stage have been previously observed in severe TBI patients^39,40^. Furthermore, the gut microbiome may be a valuable biomarker of repetitive exposure to concussions in athletes. It has been shown that concussion can trigger long-term delayed changes in the colon and that subsequent bacterial infections in the gastrointestinal tract can increase post-traumatic brain inflammation and neurodegeneration in mice^41^. Our previous studies have established a relationship between TBI and ensuing changes in the gut microbiome of mice^33^, providing the potential for a direct connection between brain-gut-microbiota and TBI. This raises the obvious question of whether the same phenomenon occurs in humans, although collecting data appropriate to test this hypothesis. We tested this hypothesis in our study by following a cohort of high-risk subjects through multiple timepoints, which yields useful. This problem has been addressed here by following a cohort of high-risk subjects through multiple timepoints, which yields useful longitudinal data from a homogenous population at the expense of comparatively low sample size among cases (in this case, *n*=4). We observe noticeable and, in some cases, statistically significant differences in the microbiome of subjects according to a concussion status and across time among those without a diagnosed concussion. These two effects, timepoint, and concussion status warrant individual discussion.

### Time-point Comparison

Although the primary hypothesis motivating this study is the effect of a diagnosed concussion on the human microbiome in the 24-48 hours following the event, a secondary hypothesis is that the chronic sub-concussive impact experienced by most football players has a comparable but less pronounced effect which subsides sometime after the season. Although not statistically significant in the classical sense, the ANOSIM analysis discussed above suggests that there may be a material change in the gut microbiome for this group happening between the post-season and off-season. The comparison between timepoints for these athletes is shown in Fig. 2. For this group, the study design uses the homogenous nature of the cohort and multiple timepoints to boost its statistical power. Still, it naturally implies that differences between timepoints (particularly in and out of season) are associated with a substantial number of common environmental changes beyond frequent collisions. We cannot associate this change with a single factor in the absence of a well-chosen control group.

One thing we can examine are specific microorganisms that show significant per-sample change in abundance based on a linear random-effects model. These microbes are presented in Supplemental Table 1, which includes the model-indicated fold-change in abundance from the post-season to the off-season. The Supplemental Table 1 is sorted according to p-value after multiple-hypothesis correction, with a cutoff of p<0.125, and includes a note about the organism’s known environment. Two observations are possible from this table. First, all the fold-change values are negative. The difference in communities is characterized by a common set of organisms that declined in abundance, but not by a common set of organisms that increased to make up the difference. In other words, the samples diverged. This is not particularly surprising since the athlete’s behavior, schedule, diet, and physical location likely also diverged as they transitioned from the rigidly structured football season to their schedules, habits, diets, and locations in the off-season.

Second, the known environment of these specific microbes suggests a possible narrative of the change. Of those with decreased average abundance, three are commonly known environmental pathogens (one of which, *S. aureus*, is known to be higher risk specifically among football teams) and the other 14 are primarily associated with soil, water, or both. All but one are under 0.5% average abundance in the postseason samples. Given these associations and low abundances the most likely explanation seems to be that these are microbes the players are exposed to regularly in the football environment, are transmitted to the gut inadvertently during the season, represent environmental contaminants, or potential misclassifications. The Fig. 2d shows the per-subject abundance change for each bacterium; the panel on the right shows that the post-to-off change for this set of bacteria appears to be highly correlated across players. Namely, about half the subjects show a clear drop in abundance across all these organisms, while the other half show small changes with no particular pattern. An interesting question for future research would be whether these players are getting some of their microbiome from the football field during the season.

### Concussion-Status Comparison

For the four players who were diagnosed with a concussion during the season, both alpha and beta diversity analyses show a significant difference between the post-concussion microbial community and that of other players during the season. The difference appears to be present in the off-season samples as well. This study is underpowered for this analysis due to the small number of cases and the fact a random-effects model is not available. Nonetheless, the comparison of subjects by concussion status and timepoint is shown in Fig. 3. The differences by concussion status are visible even in the simple stacked bar charts in Fig. 3c.

Beyond the alpha and beta diversity comparison, the multivariate analysis indicated what species are contributing most to these differences. As discussed above, the in-season model did not yield any statistically significant alterations, although the off-season model gave 9 with a p-value below 0.05, listed in Supplemental Table 2. The table also includes the coefficients and p-values for the same organisms from the in-season model, and it is worth noting that the in- and off-season coefficients are generally within a standard error of each other. It is important to re-emphasize the low statistical power in this analysis because the multiple-hypothesis correction reduces it even further. Given the difficulty of gathering a fecal sample immediately following a TBI event, the data should be considered valuable but taken lightly.

Two bacterial species were significantly depleted in relative abundance in the gut microbiota from players diagnosed with a concussion. One of the organisms identified is *Eubacterium rectale*, which was lower relative abundance (q<0.05) in the post-concussion samples. *Eubacterium rectale* is a ubiquitous member of the human gut microbiome and an anti-inflammatory microbe ^42^. A decrease in *Eubacterium rectale* has been previously correlated with peripheral inflammation in patients suffering from cognitive impairment and brain amyloidosis^43^. Also, *Eubacterium rectale* was found less frequently in children with neurodevelopmental disorders such as autism^44^. The second bacterium, *Anaerostipes hadrus*, which is a common member of the healthy human gut microbiome with known anti-inflammatory properties ^45^, showed similar depletion in the post-concussion group. Together, they hint at a narrative of the injury triggering pro-inflammatory pathways leading to a depletion in these two microbes. This is constant with effects observed in mice and underscores the need for a much larger study in the future with a large enough sample size that can provide more robust conclusions.

Next, we will review the additional results presented in the study: (i) biomarker analysis, (ii) pathway analysis, and (iii) salivary microbiome analysis.

#### (i) Biomarker analysis

There are two main limitations to the development of reliable blood TBI biomarkers: 1) Highly expressed proteins within the CNS are only detectable at low plasma concentrations, and 2) Potential biomarkers undergo proteolytic degradation in the blood, such that their levels are affected by elimination through the liver or kidney^14^. Here, we used ELISA kits for blood protein detection, in addition to the ultrasensitive multiplex array (Simoa)^15^. S100β has been proposed as a fluid biomarker following brain injury. Blood levels of S100β are elevated after mTBI and have been associated with the presence of intracranial lesions, which lead to clinical studies for the use of this biomarker as a predictor of computed tomography (CT) scan findings ^46,47^. As for SRC, serum S100β levels have been reported to be increased in athletes at 3 hours following the injury but went back to baseline levels within 2 days post-concussion ^48^. A more recent study provided evidence of elevated S100β levels at 6 hours after a concussion followed by a recovery to preseason levels in the 24-48 hours window^49^. High serum levels of SAA have been detected in children with mild head trauma ^50^, infants with white matter injury ^51^, neonates with hypoxic-ischemic encephalopathy ^52^ and adult TBI patients ^53^. However, we did not find changes in serum S100β levels across the sports season or the post-concussion athletes compared to their non-concussed teammates. But notably, both serum blood biomarkers, S100β and SAA, were positively correlated with the abundance of the *Eubacterium rectale* species. Therefore, we report that there may be a relationship between inflammatory markers in the blood and the abundance of this bacterium among football players over the course of the year for athletes experiencing repetitive subconcussive impacts. A limitation of this analysis is the high degree of variability between the biomarkers, especially for S100β, SAA, and GFAP. We could not structure the limited group of players by the position they occupy to see if this can account for the variability observed. Future studies with a larger number of participants will include the risk of receiving concussions and subconcussive hits depending on the position (e.g., lower risk for non-contact positions such as Kicker or players that do not play all the games) as a variable to be evaluated.

#### ii) Pathway analysis

Prediction of the pathway abundances associated with the fecal microbiota changes from the post to off-season indicated that processes related to the synthesis and degradation of sugars and aromatic compounds were among those showing the most significant fold changes. Interestingly, both bacteria species significantly decreased in abundance in players diagnosed with a concussion, *E. rectale*, and *A. hadrus* are major butyrate producers, with the former associated with insulin metabolism^54^. Butyrate producers are garnering renewed attention as “next-generation” probiotics for increasing colon butyrate production and their role in countering disease-associated alterations to the microbiome ^55^. Metabolomic analysis of the fecal samples in future studies will help characterize these changes further and elucidate their role in brain damage.

#### (iii) Salivary microbiome

Interestingly, we did not identify any observable changes in salivary microbiota, dampening the potential utility of this sample type as a biomarker of concussions. Despite this, a recent large prospective observational study of non-invasive concussion biomarkers has shown changes in salivary small non-coding RNAs (sncRNAs) in saliva in rugby players diagnosed with concussion^56^. The idea of having sncRNAs identifiers that correlate with microbiome signatures could represent a complete panel of concussion profiles that should be studied. Therefore, using RNA identifiers in saliva that correlate with biomarkers of the gut microbiota could represent a complete panel of concussion profiles that should be analyzed going forward.

In summary, we report alterations to the gut microbiome in collegiate football players after a single concussion and during the in-season sports activity. This is the first study to identify changes in the relative abundance of key gut bacteria in collision sports. Our results suggest that evaluating species-specific changes in gut microbiota after concussions may represent a useful diagnostic tool. Specifically, *Eubacterium rectale* was found to be associated with serum inflammatory biomarkers, indicating that maintaining the abundances of microbes commonly found in healthy adults could have neuroprotective health benefits. A significant limitation of this study is that we were following a cohort of high-risk subjects across multiple timepoints, yielding valuable longitudinal data from a homogeneous population at the expense of a comparatively low sample size among concussion cases. Since the study has low statistical power, some observations may be considered suggestive and worthy of additional study but not conclusive. For example, we need to address the history of prior concussion in future studies, incorporate the measurement of head impacts and head impact exposure over time, a depth diet analysis, or the position they occupy in the game related to the risk of receiving more exposures to hits. These and other variables can be assumed if we increase the number of participants in future studies. We plan to expand this study to increase the cohort pool during the football season in the following years to address these limitations. Undoubtedly, future research will be required to elucidate further the interactions between the gut microbiome and brain function in collision sports athletes.

## METHODS

### Study design and participants

A total of 33 male collegiate football players ages 18-23 were voluntarily recruited to the study from the same Division I university team (Fig.1). Exclusion criteria included: 1) prior history of a neurological or medical condition that could impact brain and cognition and 2) history of previous psychiatric disorder or substance abuse. The Institutional Review Board (IRB) at Houston Methodist Hospital approved the study protocol, and written informed consent was obtained from all participants. To evaluate the time course of microbiota changes in players with a clinical diagnosis of concussion or during the in-season sports activity, samples were collected at three timepoints: mid-season (sports activities start), post-season (last sports activities), and off-season (no sports activities). Since the university’s football season went from mid-August 2019 through the end of November 2019, mid-season samples were collected in October 2019, post-season in December 2019, and off-season in February 2020. (Fig.1). Additional samples were collected within 24-48 hours following a diagnosed concussion. For this study, an SRC was defined as an injury resulting from participation in organized intercollegiate practice or completions that requires medical attention by a team-certified athletic trainer or physician following the National Collegiate Athletic Association (NCAA) injury surveillance system definition ^57^. Demographics of the study cohort are summarized in Table 1.

### Sample collection and bacterial DNA extraction

Stool samples were collected using the Stool Nucleic Acid Collection and Preservation System (Norgen, Ontario, Canada). Approximately 2 g of stool was sampled using the sterile spoon attached to the lid of the Stool Nucleic Acid Collection and Transport Tube. The sample was then suspended in the preservative solution and mixed thoroughly. Samples were kept at room temperature and, upon arrival to the laboratory, further homogenized by vortexing. They were then stored at 4 °C. For DNA extraction, 400 μl of the fecal suspension was transferred to the bead tubes supplied in the QIAamp PowerFecal Pro DNA Kit (Qiagen, Germantown, MD). Bead beating was performed for 1 min at 6.5 m/s on a FastPrep-24 system (MP Biomedicals, Irvine, CA). DNA isolation then continued following the kit manufacturer’s instructions. Saliva samples were collected as indicated in the Collection Tube. The Collection Funnel was removed, the contents of the Preservative Ampoule were added and mixed with the collected saliva. Samples were kept at room temperature and then stored at 4 °C until processing. DNA was extracted from the saliva samples using the Norgen Saliva DNA Isolation Reagent Kit (Norgen, Ontario, Canada), per the manufacturer’s instructions.

### Long-read 16S rRNA sequencing on the MinION platform

16S rRNA amplicon sequencing was performed on a MinION nanopore sequencer (Oxford Nanopore Technologies, Oxford, UK). The amplicon library was prepared using the 16S Barcoding Kit 1-24 (SQK-16S024, Oxford Nanopore Technologies, Oxford, UK). For the PCR amplification and barcoding, 15 ng of template DNA extracted from fecal samples, and 30 ng in the case of saliva, were added to the LongAmp Hot Start Taq 2X Master Mix (New England Biolabs, Ipswich, MA). Initial denaturation at 95 °C was followed by 35 cycles of 20 s at 95 °C, 30 s at 55 °C, 2 min at 65 °C, and a final extension step of 5 min at 65 °C. Purification of the barcoded amplicons was performed using the AMPure XP Beads (Beckman Coulter, Brea, CA) as per Nanopore’s instructions. Samples were then quantified using Qubit fluorometer (Life Technologies, Carlsbad, CA) and pooled in an equimolar ratio to a total of 50-100 ng in 10 μl. The pooled library was then loaded into an R9.4.1 flow cell and run per the manufacturer’s instructions. MINKNOW software 19.12.5 was used for data acquisition. The provided primer set includes a recently noted issue with the standard ONT forward primer, which contains three mismatching bases to the family *Bifidobacteriaceae* and thus fails to amplify microbes of this taxa ^58^.

### Long-read 16S rRNA sequencing data taxonomic analysis

Raw fastq sequences were base called using Guppy. Sequencing adapters were then removed from sequences with Porechop v0.2.4. The resulting reads were then classified taxonomically with Kraken v1.1.1 ^35^. The Pavian web application (https://fbreitwieser.shinyapps.io/pavian/) was utilized to transform Kraken outputs into spreadsheets of taxa counts and relative abundances per sample phylum, family,, genus, and species levels. This was completed by: 1) uploading all Kraken. kreport text files for a given sample type, 2) selecting the “Comparison” tab on the web browser, 3) selecting either “read” or “%,” the appropriate taxonomy rank, and “clade” to ensure all bacteria classified at lower taxonomy level were included in higher-level analysis, and 4) downloading the full tab-separated table. To reduce noise, any bacteria that claimed no more than 2 reads in at least one sample were removed from further analysis. Bacteria classified at the species or strain level that were missing genera or family in their lineage due to NCBI taxonomy were edited to ensure these bacteria were included in the genus- and family-level analysis. This impacted two bacterial species at the genus level: *Eubacterium rectale* and *Ruminococcaceae bacterium* CPB6. To match NCBI Taxonomy Database, their listed genera, unclassified Lachnospiraceae, and unclassified Ruminococcaceae, respectively, were treated as classified genera. This also impacted two bacterial species at the family level: *Intestinimonas butyriciproducens* and *Flavonifractor plautii*. Both bacteria claim taxonomic family *unclassified Clostridiales*, which is treated as a classified family for our analysis.

Alpha and beta diversity analyses for microbiome samples were performed with Python 3.8 using the sci-kit bio v0.4.1 diversity package. Alpha diversity values were generated for each sample using the Shannon and Simpson metrics. Beta diversity analyses were observed with the Weighted UniFrac metric. A phylogenetic tree was generated from the Kraken database using the taxdump_to_tree.py python script in biocore, a space for collaboratively developed bioinformatics software. Each branch in the tree was given a length of 1. Beta diversity distance matrices were then generated for each of the two analysis workflows per sample type. To evaluate the statistical difference between groups, p-values were calculated for the entire workflow and each pairwise grouping. This was completed by simulating 9999 ANOSIM permutations for each analysis. To visualize these values, a Matplotlib v.3.3.3 beta diversity scatter plot was generated for each workflow. This was achieved by: generating a principal coordinate analysis table from the appropriate distance matrix, choosing axes that best represented the calculated p-values, and plotting points colored by group. Finally, 95% confidence ellipses were drawn around each group with Matplotlib two-dimensional confidence ellipse source code.

Stacked bar plots were created with GraphPad Prism version 9 to visualize the average relative abundance of bacteria amongst samples in each group for the given workflow. These values were calculated at the phylum, family, and genus level by averaging the appropriate samples using the aligned relative abundance spreadsheet described above. Log2 fold change was calculated between two samples from the same participant by taking the log2 of the percent relative abundance of the later timepoint divided by an earlier timepoint. To avoid mathematical errors, relative abundance values of 0 were transformed to 10^−10^. Finally, heatmaps were created with the heatmap function in Seaborn v0.11.0 with the calculated fold change values. Scripts for generating diversity plots and heatmaps can be found here: https://gitlab.com/treangenlab/tbi-microbiome-football-2019.

### Prediction of pathway composition by PICRUSt2

Metagenomic functional composition from the taxa abundance was inferred using PICRUSt2^36^. An amplicon sequence variant (ASV) table was generated where the taxonomy of each ASV was assigned by Kraken 2 classification against the Silva database version 138^35^. Abundance of MetaCyc pathways was then estimated using PICRUSt2 with default options. Pathways with relative abundance < 0.01 % were converted to 0 to reduce noise.

### Measurement of serum inflammatory biomarkers

To accurately measure the candidate blood biomarkers, ELISA kits and SIMOA Ultrasensitive assays were performed. Venous blood was collected into vacutainer sterile serum tubes and allowed to clot at room temperature for 30 min. After centrifugation at 3000 g for 15 min, serum was aliquoted and stored at -80 °C until analysis. Serum NF-L, GFAP, total Tau, and ubiquitin carboxy-terminal hydrolase L1 (UCH-L1) were quantified using the SIMOA Neurology 4-plex assay. According to the manufacturer’s protocol, samples were run in duplicates on a HD-X instrument (Quanterix, Lexington, MA). We did not find any differences with Tau and UCH-L1 markers. The assay range was calculated from the calibration curve for each of the analytes. The functional lower limit of quantification was estimated at 1.052 pg/mL for NF-L, 1.728 pg/mL for GFAP, 0.576 pg/mL for Tau, and 13.520 pg/mL for UCH-L1. According to the manufacturer’s instructions, serum levels of S100β and SAA were measured using a commercial enzyme-linked immunosorbent assay (ELISA) to verify previous SIMOA data according to the manufacturer’s instructions (Human S100B, EZHS100B-33K, Millipore Sigma, Burlington, MA; Human SAA, EL10015L, Anogen, Mississauga, Canada). The lower limit of detection for the S100β and SAA assays were 2.7 pg/mL and 1.1 ng/mL, respectively. When S100β was not detectable, the limit of detection value was assigned to the sample. Each sample was measured in duplicate.

### Optic nerve sheath diameter and velocities with breath-holding

Each participant’s optic nerve sheath diameter (ONSD) was measured using a transcranial Doppler (TCD). ONSDs were measured during the post-season and off-season periods. A high-frequency General Electric (Fairfield, CT, US) LOGIQ E9 9 MHz linear probe was utilized for this study. All measurements were performed by a single, experienced, certified neuro-sonographer using the same ultrasound machine to eliminate any operator bias. The probe was placed over the closed eyelid with the patient in supine position. A small amount of ultrasound gel was used to avoid any discomfort for the patient. The probe was held in a transverse plane with the beam directed posteriorly towards the optic disc complex. The patient was asked to look down while keeping their eyes closed. The ONSD was measured 3 mm behind the globe using an electronic caliper. Each diameter was measured three times, and the mean value was used for the final analysis. Velocities were measured in corresponding locations with temporal (ACA, MCA, PCA), transorbital (ONSD and OA), and suboccipital (VA and BA) windows. BHI and delta velocities were calculated utilizing a 20 second player-patient initiated inhalation and hold.

### Statistical analysis

Statistical analysis of the alpha diversity in the microbiome and serum biomarkers was performed using GraphPad Prism version 9. A Kruskal-Wallis test followed by Dunn’s *post hoc* test for multiple comparisons was performed for the Shannon and Simpson alpha diversity indices. Repeated measures one-way ANOVA was used for the longitudinal analysis of the serum biomarkers, whereas an ordinary one-way ANOVA was performed to find statistical differences between the post-concussion samples and the rest of the groups.

To analyze whether any individual species exhibited a significant difference in relative abundance between timepoints or across health status, we constructed linear models in MaAsLin2 (MaAsLin2 R package version 1.4.0, http://huttenhower.sph.harvard.edu/maaslin2). For all models, relative abundance data was calculated for each microbe as a percentage of the total sample, and default transformation, normalization, and outcome distribution options were selected. For the longitudinal analysis among healthy players (n = 17 players x 3 sample times), a random-effects model was selected with the player as the random effect and the fixed effect was specified to be the timepoint (mid/post/off-season); features (i.e., species) were only modeled if they were present with a minimum abundance of 0.05% in at least 10% of the samples (i.e., 6 or more). For the concussion-vs. non-concussed samples, models were run separately for in-season (i.e., mid and post-season, combined, n = 51 healthy, 4 concussion) and off-season (n = 21 healthy, 4 concussion). For both models, concussion status was specified as the fixed effect and features were only included if they were present with a relative abundance of at least 1% in a minimum of 10% of the samples. In this case, the higher abundance threshold was due to the small number of cases available, to reduce unnecessary hypothesis tests requiring correction.

To examine the interactions between the serum biomarkers and the gut microbiome composition, Pearson correlations were computed between the concentration of biomarkers throughout the sports season in non-concussed athletes and the relative abundance of species and genus found to be significantly changed in the gut microbiome analyses. Pearson correlation tests followed by false discovery rate (FDR) adjustment were performed in GraphPad v9.

## Data Availability

Data availability. The high-throughput sequence data has been deposited in the National Center for Biotechnology Information (NCBI) Sequence Read Archive (SRA) under project database with project number PRJNA699316. The authors declare that all other data supporting the findings of this study are available within the article and its Supplementary Information files, or from the corresponding author on request.

## Data availability

The high-throughput sequence data has been deposited in the National Center for Biotechnology Information (NCBI) Sequence Read Archive (SRA) under the project database with project number PRJNA699316. The authors declare that all other data supporting the findings of this study are available within the article and its Supplementary Information files or from the corresponding author on request.

## Acknowledgments

This work was supported by grants from the National Institute for Neurological Disorders and Stroke (NINDS) R21NS106640 (SV) and funds from Houston Methodist Research Institute (SV, TT). The authors are indebted to Dr. Gillian Hamilton for editing. Figure 1 was created using Biorender.com.

## Author Contributions

S.V., S.Sa., R.G., K.P., G.B., T.T., and S.Sc. designed the study, S.Sa., A.C., and S.Sc. coordinated the participants enrollment, J.W., A.C., S.So. collected the samples, R.K. performed TCD analysis, S.So. performed laboratory analysis, K.C., Q.W., M.N., E.R., and T.T. performed bioinformatic and statistical analysis, and S.V., T.T., S.So., K.C., and M.N. wrote the manuscript, S.V., T.T., R.G., S.So., K.C., and M.N. analyzed the data, S.V. and T.T. coordinated the study. All authors reviewed and revised the final manuscript.

## REFERENCES

1 Howell, D. R., Kirkwood, M. W., Laker, S. & Wilson, J. C. Collision and Contact Sport Participation and Quality of Life Among Adolescent Athletes. J Athl Train 55, 1174–1180, doi:10.4085/1062-6050-0536.19 (2020).

2 McCrory, P. et al. Consensus statement on Concussion in Sport - The 4th International Conference on Concussion in Sport held in Zurich, November 2012. Phys Ther Sport 14, e1–e13, doi:10.1016/j.ptsp.2013.03.002 (2013).

3 Li, A. Y. et al. Sport Contact Level Affects Post-Concussion Neurocognitive Performance in Young Athletes. Arch Clin Neuropsychol, doi:10.1093/arclin/acab021 (2021).

4 Conder, R. L. & Conder, A. A.Sports-related concussions. N C Med J 76, 89–95, doi:10.18043/ncm.76.2.89 (2015).

5 Jordan, B. D. The clinical spectrum of sport-related traumatic brain injury. Nat Rev Neurol 9, 222–230, doi:10.1038/nrneurol.2013.33 (2013).

6 Shuttleworth-Edwards, A. B., Smith, I. & Radloff, S. E. Neurocognitive vulnerability amongst university rugby players versus noncontact sport controls. J Clin Exp Neuropsychol 30, 870–884, doi:10.1080/13803390701846914 (2008).

7 Iverson, G. L., Williams, M. W., Gardner, A. J. & Terry, D. P. Systematic Review of Preinjury Mental Health Problems as a Vulnerability Factor for Worse Outcome After Sport-Related Concussion. Orthop J Sports Med 8, 2325967120950682, doi:10.1177/2325967120950682 (2020).

8 Reynolds, B. B. et al. Practice type effects on head impact in collegiate football. J Neurosurg 124, 501–510, doi:10.3171/2015.5.JNS15573 (2016).

9 Duma, S. M. et al. Analysis of real-time head accelerations in collegiate football players. Clin J Sport Med 15, 3–8, doi:10.1097/00042752-200501000-00002 (2005).

10 Guskiewicz, K. M. et al. Measurement of head impacts in collegiate football players: relationship between head impact biomechanics and acute clinical outcome after concussion. Neurosurgery 61, 1244-1252; discussion 1252-1243, doi:10.1227/01.neu.0000306103.68635.1a (2007).

11 Manley, G. et al. A systematic review of potential long-term effects of sport-related concussion. Br J Sports Med 51, 969–977, doi:10.1136/bjsports-2017-097791 (2017).

12 Montenigro, P. H. et al. Cumulative Head Impact Exposure Predicts Later-Life Depression, Apathy, Executive Dysfunction, and Cognitive Impairment in Former High School and College Football Players. J Neurotrauma 34, 328–340, doi:10.1089/neu.2016.4413 (2017).

13 Mouzon, B. C. et al. Lifelong behavioral and neuropathological consequences of repetitive mild traumatic brain injury. Ann Clin Transl Neurol 5, 64–80, doi:10.1002/acn3.510 (2018).

14 Gallo, V. et al. Concussion and long-term cognitive impairment among professional or elite sport-persons: a systematic review. J Neurol Neurosurg Psychiatry 91, 455–468, doi:10.1136/jnnp-2019-321170 (2020).

15 Stein, T. D., Alvarez, V. E. & McKee, A. C. Concussion in Chronic Traumatic Encephalopathy. Curr Pain Headache Rep 19, 47, doi:10.1007/s11916-015-0522-z (2015).

16 Smoliga, J. M. Interpreting Biomarker Data After Concussion and Repeated Subconcussive Head Impacts: Challenges in Evaluating Brain Protection. JAMA Neurol 77, 1477–1478, doi:10.1001/jamaneurol.2020.3467 (2020).

17 Oliver, J. M. et al. Fluctuations in blood biomarkers of head trauma in NCAA football athletes over the course of a season. J Neurosurg, 1–8, doi:10.3171/2017.12.JNS172035 (2018).

18 Harmon, K. G. et al. American Medical Society for Sports Medicine Position Statement on Concussion in Sport. Clin J Sport Med 29, 87–100, doi:10.1097/JSM.0000000000000720 (2019).

19 Jindal, G., Gadhia, R. R. & Dubey, P. Neuroimaging in Sports-Related Concussion. Clin Sports Med 40, 111–121, doi:10.1016/j.csm.2020.08.004 (2021).

20 Zhang, J. et al. Relationship between white matter integrity and post-traumatic cognitive deficits: a systematic review and meta-analysis. J Neurol Neurosurg Psychiatry 90, 98–107, doi:10.1136/jnnp-2017-317691 (2019).

21 Wallace, E. J., Mathias, J. L. & Ward, L. Diffusion tensor imaging changes following mild, moderate and severe adult traumatic brain injury: a meta-analysis. Brain Imaging Behav 12, 1607–1621, doi:10.1007/s11682-018-9823-2 (2018).

22 Mondello, S. et al. Blood-based diagnostics of traumatic brain injuries. Expert Rev Mol Diagn 11, 65–78, doi:10.1586/erm.10.104 (2011).

23 Di Battista, A. P., Churchill, N., Rhind, S. G., Richards, D. & Hutchison, M. G. Evidence of a distinct peripheral inflammatory profile in sport-related concussion. J Neuroinflammation 16, 17, doi:10.1186/s12974-019-1402-y (2019).

24 Mazarati, A., Medel-Matus, J. S., Shin, D., Jacobs, J. P. & Sankar, R. Disruption of intestinal barrier and endotoxemia after traumatic brain injury: Implications for post-traumatic epilepsy. Epilepsia, doi:10.1111/epi.16909 (2021).

25 Grande, P. O., Asgeirsson, B. & Nordstrom, C. Aspects on the cerebral perfusion pressure during therapy of a traumatic head injury. Acta Anaesthesiol Scand Suppl 110, 36–40, doi:10.1111/j.1399-6576.1997.tb05493.x (1997).

26 Benakis, C. et al. Commensal microbiota affects ischemic stroke outcome by regulating intestinal gammadelta T cells. Nat Med 22, 516–523, doi:10.1038/nm.4068 (2016).

27 Tan, M., Zhu, J. C., Du, J., Zhang, L. M. & Yin, H. H. Effects of probiotics on serum levels of Th1/Th2 cytokine and clinical outcomes in severe traumatic brain-injured patients: a prospective randomized pilot study. Crit Care 15, R290, doi:10.1186/cc10579 (2011).

28 Brenner, L. A. et al. Growing literature but limited evidence: A systematic review regarding prebiotic and probiotic interventions for those with traumatic brain injury and/or posttraumatic stress disorder. Brain Behav Immun 65, 57–67, doi:10.1016/j.bbi.2017.06.003 (2017).

29 Cenit, M. C., Sanz, Y. & Codoner-Franch, P. Influence of gut microbiota on neuropsychiatric disorders. World J Gastroenterol 23, 5486–5498, doi:10.3748/wjg.v23.i30.5486 (2017).

30 Arciniegas, D. B. & McAllister, T. W. Neurobehavioral management of traumatic brain injury in the critical care setting. Crit Care Clin 24, 737-765, viii, doi:10.1016/j.ccc.2008.06.001 (2008).

31 McAllister, T. W. Neurobehavioral sequelae of traumatic brain injury: evaluation and management. World Psychiatry 7, 3–10, doi:10.1002/j.2051-5545.2008.tb00139.x (2008).

32 Hoban, A. E. et al. The microbiome regulates amygdala-dependent fear recall. Mol Psychiatry 23, 1134–1144, doi:10.1038/mp.2017.100 (2018).

33 Treangen, T. J., Wagner, J., Burns, M. P. & Villapol, S. Traumatic Brain Injury in Mice Induces Acute Bacterial Dysbiosis Within the Fecal Microbiome. Front Immunol 9, 2757, doi:10.3389/fimmu.2018.02757 (2018).

34 Nygaard, A. B., Tunsjo, H. S., Meisal, R. & Charnock, C. A preliminary study on the potential of Nanopore MinION and Illumina MiSeq 16S rRNA gene sequencing to characterize building-dust microbiomes. Sci Rep 10, 3209, doi:10.1038/s41598-020-59771-0 (2020).

35 Wood, D. E., Lu, J. & Langmead, B. Improved metagenomic analysis with Kraken 2. Genome Biol 20, 257, doi:10.1186/s13059-019-1891-0 (2019).

36 Douglas, G. M. et al. PICRUSt2 for prediction of metagenome functions. Nat Biotechnol 38, 685–688, doi:10.1038/s41587-020-0548-6 (2020).

37 Han, M. et al. Stratification of athletes’ gut microbiota: the multifaceted hubs associated with dietary factors, physical characteristics and performance. Gut Microbes 12, 1–18, doi:10.1080/19490976.2020.1842991 (2020).

38 Ribeiro, F. M., Petriz, B., Marques, G., Kamilla, L. H. & Franco, O. L. Is There an Exercise-Intensity Threshold Capable of Avoiding the Leaky Gut? Front Nutr 8, 627289, doi:10.3389/fnut.2021.627289 (2021).

39 Urban, R. J. et al. Altered Fecal Microbiome Years after Traumatic Brain Injury. J Neurotrauma 37, 1037–1051, doi:10.1089/neu.2019.6688 (2020).

40 Nicholson, S. E. et al. Moderate Traumatic Brain Injury Alters the Gastrointestinal Microbiome in a Time-Dependent Manner. Shock 52, 240–248, doi:10.1097/SHK.0000000000001211 (2019).

41 Ma, E. L. et al. Bidirectional brain-gut interactions and chronic pathological changes after traumatic brain injury in mice. Brain Behav Immun 66, 56–69, doi:10.1016/j.bbi.2017.06.018 (2017).

42 Mukherjee, A., Lordan, C., Ross, R. P. & Cotter, P. D. Gut microbes from the phylogenetically diverse genus Eubacterium and their various contributions to gut health. Gut Microbes 12, 1802866, doi:10.1080/19490976.2020.1802866 (2020).

43 Cattaneo, A. et al. Association of brain amyloidosis with pro-inflammatory gut bacterial taxa and peripheral inflammation markers in cognitively impaired elderly. Neurobiol Aging 49, 60–68, doi:10.1016/j.neurobiolaging.2016.08.019 (2017).

44 Bojovic, K. et al. Gut Microbiota Dysbiosis Associated With Altered Production of Short Chain Fatty Acids in Children With Neurodevelopmental Disorders. Front Cell Infect Microbiol 10, 223, doi:10.3389/fcimb.2020.00223 (2020).

45 Brereton, N. J. B., Pitre, F. E. & Gonzalez, E. Reanalysis of the Mars500 experiment reveals common gut microbiome alterations in astronauts induced by long-duration confinement. Comput Struct Biotechnol J 19, 2223–2235, doi:10.1016/j.csbj.2021.03.040 (2021).

46 Bazarian, J. J. et al. Classification accuracy of serum Apo A-I and S100B for the diagnosis of mild traumatic brain injury and prediction of abnormal initial head computed tomography scan. J Neurotrauma 30, 1747–1754, doi:10.1089/neu.2013.2853 (2013).

47 Zongo, D. et al. S100-B protein as a screening tool for the early assessment of minor head injury. Ann Emerg Med 59, 209–218, doi:10.1016/j.annemergmed.2011.07.027 (2012).

48 Kiechle, K. et al. Subject-specific increases in serum S-100B distinguish sports-related concussion from sports-related exertion. PLoS One 9, e84977, doi:10.1371/journal.pone.0084977 (2014).

49 Meier, T. B. et al. Prospective Assessment of Acute Blood Markers of Brain Injury in Sport-Related Concussion. J Neurotrauma 34, 3134–3142, doi:10.1089/neu.2017.5046 (2017).

50 Gao, X. et al. A “hot Spot”-Enhanced paper lateral flow assay for ultrasensitive detection of traumatic brain injury biomarker S-100beta in blood plasma. Biosens Bioelectron 177, 112967, doi:10.1016/j.bios.2021.112967 (2021).

51 Leviton, A. et al. Early postnatal blood concentrations of inflammation-related proteins and microcephaly two years later in infants born before the 28th post-menstrual week. Early Hum Dev 87, 325–330, doi:10.1016/j.earlhumdev.2011.01.043 (2011).

52 Aly, H. et al. Serum amyloid A protein and hypoxic ischemic encephalopathy in the newborn. J Perinatol 31, 263–268, doi:10.1038/jp.2010.130 (2011).

53 Hergenroeder, G. et al. Identification of serum biomarkers in brain-injured adults: potential for predicting elevated intracranial pressure. J Neurotrauma 25, 79–93, doi:10.1089/neu.2007.0386 (2008).

54 Venkataraman, A. et al. Variable responses of human microbiomes to dietary supplementation with resistant starch. Microbiome 4, 33, doi:10.1186/s40168-016-0178-x (2016).

55 Boesmans, L. et al. Butyrate Producers as Potential Next-Generation Probiotics: Safety Assessment of the Administration of Butyricicoccus pullicaecorum to Healthy Volunteers. mSystems 3, doi:10.1128/mSystems.00094-18 (2018).

56 Di Pietro, V. et al. Unique diagnostic signatures of concussion in the saliva of male athletes: the Study of Concussion in Rugby Union through MicroRNAs (SCRUM). Br J Sports Med, doi:10.1136/bjsports-2020-103274 (2021).

57 Kilcoyne, K. G. et al. Reported Concussion Rates for Three Division I Football Programs: An Evaluation of the New NCAA Concussion Policy. Sports Health 6, 402–405, doi:10.1177/1941738113491545 (2014).

58 Fujiyoshi, S., Muto-Fujita, A. & Maruyama, F. Evaluation of PCR conditions for characterizing bacterial communities with full-length 16S rRNA genes using a portable nanopore sequencer. Sci Rep 10, 12580, doi:10.1038/s41598-020-69450-9 (2020).

